# The Impact of Self-Reported Health Factors on Behavioural Difficulties in School-Aged Children: Findings from the HAPPEN Pan-Wales Cohort Using Data Linkage

**DOI:** 10.64898/2026.07.25.26358910

**Authors:** Michaela James, Amy Locke, Jon Kennedy, Mayara Silveira Bianchim, Sam Dredge, Liam Mahedy, Sinead Brophy

## Abstract

This study aimed to examine factors associated with behavioural difficulties among school-aged children (aged 7 -11 years) using self-reported survey data linked to education data from a national cohort study of over 50,000 children (the HAPPEN national cohort) in Wales, UK, between 2014 and 2024.

Data include self-reported health and wellbeing linked to Additional Learning Needs (ALN) status collected from education data in Wales. Measures included self-reported time spent on screens, frequency of sugary snack, fizzy drink and takeaway food consumption, physical activity levels, sleep duration and behavioural difficulties (as measured by the Me and My Feelings Survey). Associations were analysed using descriptive trends, K-means cluster analysis and multivariate regression modelling.

Findings showed a rise in behavioural difficulties during and after the Covid-19 pandemic, which are now improving in Wales. Cluster analysis identified that children with poor sleep and high consumption of takeaway foods and fizzy drinks dietary habits had the highest levels of behavioural difficulties, a trend that remained significant after adjusting for all potential confounders. Physical activity associated with reduced behavioural difficulties, with 5–6 days of activity associated with the lowest difficulty scores.

These findings suggest that behavioural difficulty is driven by health behaviours especially sleep and diet rather than single factors. The results highlight the need for integrated public health approaches that address dietary quality and sleep hygiene rather than focusing on individual behaviours in isolation. Promoting balanced daily activity patterns, establishing good sleep routines, improving food environments and building movement into the school day may have important benefits for children’s behaviour.

## Introduction

Behavioural difficulties (e.g., angry outbursts, hitting objects, distancing socially, or ignoring rules) in classrooms represent a significant and growing challenge, there remains a gap in the literature regarding interventions (1). There is increasing concern about the rising number of children being suspended from school, including pre-school and primary school pupils, as a behaviour management strategy (2). While low intensity (‘naughty’) and impulsive behaviour may be regarded as a normal part of child development, more persistent and challenging behaviours outside of those expected for a child’s age and developmental stage can be interpreted as a lack of coping strategies, additional learning needs or impaired social and communication skills (3).

Several risk factors and precursors for behavioural difficulties have been identified (1). Children are growing up in spaces where technology has become an integral part of their lives (4). While television viewing remains the most common type of screen-based activity, video gaming, computer and laptop use, and ownership of tablets and smartphones have become increasingly prevalent and commonplace for young children (5,6). The widespread availability and declining cost of technology has contributed to this trend (7) and a recent report from the House of Commons (2024) estimated that around half of children and young people are exceeding the public health recommendations of no more than two hours of recreational screen time per day or less (8). The World Health Organisation (WHO) recommends no more than one hour a day of sedentary screen time for children aged 2-4 years (9).

Emerging research has highlighted associations between being greater sedentary behaviour, increased screen time and poorer mental health in young people (10,11). More specifically, growing evidence suggests that increased screen time use is associated with behavioural difficulties in children (6,7,12–16). These behaviours may have important implications for children’s relationships, their school life and beyond (17). Children who have challenging behaviours are at risk of academic failure, dropping out of school and risky behaviour engagement (2).

Lifestyle factors such as increased screen time and poor dietary habits have become central concerns for children’s general health and wellbeing. Over recent decades, the time children spend using electronic devices has been increasing significantly (6,18). Sedentary time has well established associations with cardiometabolic disorders, including obesity and high blood pressure as well as sleep disorders (4,8,19,20). These associations are partly attributable to sustained periods of inactivity and increased links with the consumption of energy-dense foods (19). Several studies highlight that prolonged use of screens correlates with unhealthier dietary habits, such as meal skipping, increased snacking, binge eating, higher consumption of energy dense foods and lower fruit and vegetable intake (5,21–23). These lifestyle behaviours have also been linked to wider implications around behaviour, attention problems and attainment in school (6,10,14,23).

Other health behaviours including physical activity (PA) and quantity of sleep, are also important contributors to behaviour in children. Regular PA has positive effects on cognitive function, attention and cognitive performance in children (24,25). Research has shown it is a potential strategy for increasing behavioural engagement in the classroom (26). Insufficient sleep and poor sleep quality have been shown to negatively affect cognitive function in children, including attention and academic performance (27). Research has demonstrated links between sleep duration and behavioural difficulties, including acting out behaviours (28,29).

This work aimed to examine the relationship between key, self-reported health behaviours (screen use, nutrition, physical activity and sleep) and behavioural difficulties in primary school-aged children (7-11 years) in Wales using data from HAPPEN (Health and Attainment of Pupils in Primary Education) Cohort (30). The findings from this work can help better inform policy and practice around recommendations for good and for harm when discussing behaviour in children. This study was formulated following conversations with key interest groups in the children’s health and wellbeing space (e.g., practitioners, educators and local and national government representatives) whereby concerns were raised around the impact of screen use on behaviour in school. This study addresses a gap in the evidence on effective school-based interventions and responds to concerns raised by key contributors and interested parties.

## Materials and Methods

Data were taken from the Health and Attainment of Pupils in Primary Education (HAPPEN) cohort (30). This is a pan-Wales cohort of over 50,000 children aged 7-11 which aims to empower schools by bringing together education, health and research in line with the Curriculum for Wales (CfW) where Health and Wellbeing is a core aspect of the six Areas of Learning Experience (31). The survey was co-developed following interviews with headteachers who called for a better understanding of pupils needs (32,33). The study uses data from 2014 to 2024. It aims to investigate whether screen time (defined as 2+ hours per day) and sugary snack consumption influences the behavioural difficulties of school-aged children. Using data from HAPPEN linked with additional learning needs (ALN) diagnosis in the Secure Anonymised Information Linkage (SAIL) Databank (34) allows this study to provide a novel approach of exploring outcomes linked with behavioural difficulties as reported by the children themselves. For this study, ALN was taken from the EDUW dataset which includes diagnosis across several ALN indicators including ADHD, autism and physical disabilities. Specifically, the research seeks to:

1. Identify trends in self-reported sedentary behaviour, sugary snack intake and behavioural outcomes across different gender groups (boy, girl, prefer not to say) from 2014 to 2024.
2. Explore the associations between these variables and how health behaviours cluster together.

### Participants

Schools take part in HAPPEN on a voluntary basis, with recruitment opening every September in line with the beginning of the new school term in Wales. Targeted recruitment takes the form of direct communication with schools within the HAPPEN network and wider recruitment takes place through key interest holder engagement via local authority education teams, community development teams, charities and word of mouth. Further detail about recruitment can be found in HAPPEN’s Cohort Profile paper (30). As of 2025, over 650 schools have registered to take part in HAPPEN.

### Patient and Public Involvement

Patient and public involvement (PPI) is integral to the growth and purpose of HAPPEN, underscoring its commitment to participatory and proactive research that aims to empower schools. Valuable input was sought during the design of the surveys from children, parents and school staff. The inclusion of PPI ensures that interest groups are actively participate in shaping research. For this study, conversations with key interest groups including parents, teachers, child-care practitioners, social justice teams and Welsh Government officials from various engagement meetings identified the need to explore links between screen use (sedentary time), diet (specifically, sugary snacks) and behavioural difficulties.

### Data Collection

Schools can take part in the HAPPEN Survey throughout the academic year to provide snapshots, track change and evaluate practice. Having completed the survey, schools receive an individual school report aligned with the CfW presenting average data compared with national averages of health and wellbeing in the school. Once a school has decided to take part in HAPPEN, they receive an information pack and communicate with parents. Parents receive the information sheets and opt-out forms prior to the school taking part and can opt their child out of taking part. This process is governed by opt-out consent.

All pupils in years 4, 5 and 6 (ages 7–11) can complete the HAPPEN Survey; an online health and wellbeing questionnaire focused on physical and mental health. The survey asks children about their general health and wellbeing; specifically, physical activity (self-reported days with activity time equating to 2+ hours or more over 7 days) and screen time (self-reported days with sedentary time equating to 2+ hours spent using screens over 7 days), sleep, diet and dental health (takeaways, fizzy drinks), emotional and behavioural difficulties (the ‘Me and My Feelings’ survey), the Good

Childhood Index (35) and general wellbeing (autonomy and competence, happiness). A full copy of the survey can be seen as supplementary file 1.

### Data Analysis

Descriptive analysis was used to examine trends over time in screen time, diet, and behavioural difficulty across gender groups (boys, girls, and prefer not to say). Mean behavioural difficulty scores (as the continuous outcome, scoring from 0-12 with 12 being the highest possible) were reported.

K-Means clustering analysis was used to examine associations between screen time, sugary snack consumption, fizzy drinks, takeaways and behavioural difficulties. Seven behavioural variables (screen time, consumption of sugary snacks, fizzy drinks, and takeaways, physical activity, sleep duration, and behavioural difficulties) were first standardised into z-scores. This was done to prevent variables with larger scales from disproportionately influencing the cluster formation While the optimal number of clusters was determined using the Elbow method using the sum of squares (WSS), the Akaike Information Criterion (AIC), the Bayesian Information Criterion (BIC), and the Calinski-Harabasz Index (CIH) (36). To determine whether there are statistical differences between the clusters selected based on the optimal number of clusters, the homogeneity of variance was tested using the Levene test. All statistical analyses were performed using SPSS (version 19).

To explore the independent associations in more detail between self-reported health behaviours and behavioural difficulties, multivariable linear regression model was carried out. Regression coefficients are unstandardised and represent the adjusted difference in mean behavioural difficulty score compared with the reference category, holding the other variables in the model constant. The continuous outcome variable was the behavioural difficulty score. Predictor variables in the model included demographic characteristics; weekly health behaviours and controlling for ALN status (No ALN vs. ALN) as a confounder. alongside weekly health behaviours. Children with ALN are more likely to experience behavioural difficulties and may also differ in their health behaviours. Inclusion of ALN status in the analyses enabled an assessment of whether associations between health behaviours and behavioural difficulties were independent of ALN status.

After the clusters were established, the distribution of participants across groups was recorded, and the mean z-scores for each input variable were calculated by cluster membership to facilitate the interpretation and labelling of the resulting behavioural profiles.

## Results

A total of 55,741 responses were included in the analysis dating from between 2014 and 2024. This was split as 44 % boys (n=24,526), 48 % girls (n=26,756) and 2 % preferring not to disclose their gender (n=1,115); 6 % of data for gender were missing (n=3344). A demographic breakdown in relation to behavioural difficulties reported can be seen as Table 1.

**Table 1.** Demographics of Cohort.

|  | **Clinical**<br>**Behavioural**<br>**Difficulties**<br>**(n=3,961)** | **Borderline**<br>**Behavioural**<br>**Difficulties**<br>**(n=2,902)** | **No**<br>**Behavioural**<br>**Difficulties**<br>**(n=43,549)** |
| --- | --- | --- | --- |
| **Gender** |  |  |  |
| Boys | 56 % (n=2,213) | 59 % (n=1,713) | 45 % (n=19,546) |
| Girls | 38 % (n=1,518) | 36 % (n=1,057) | 53 % (n=22,938) |
| Prefer Not To Say | 6 % (n=188) | 5 % (n=111) | 2 % (n=1,223) |
| Missing | 1 % (n=42) | 1 % (n=21) | . |
| **Additional Learning Needs** |  |  |  |
| No ALN | 65 % (n=2,586) | 68 % (n=1,965) | 76 % (n=33,041) |
| ALN | 33 % (n=1,312) | 31 % (n=886) | 22 % (n=9,712) |
| Missing | 2 % (n=63) | 1 % (n=51) | 2 % (n=796) |
| **Screen Time (How many days do you spend watching a screen for 2+ hours?)** |  |  |  |
| 0 days | 4 % (n=146) | 3 % (n=86) | 4 % (n=1,875) |
| 1-2 days | 14 % (n=557) | 15 % (n=437) | 20 % (n=8,776) |
| 3-4 days | 14 % (n=535) | 28 % (n=810) | 20 % (n=8,873) |
| 5-6 days | 13 % (n=496) | 12 % (n=354) | 3 % (n=1,325) |
| 7 days | 54 % (n=2,155) | 42 % (n=1,222) | 38 % (n=16,540) |
| Missing | 2 % (n=72) | . | 14 % (n=6,160) |
| **Sugary Snacks (How many days did you eat at least one sugary snack [e.g. chocolate bar, sweets]?)** |  |  |  |
| 0 days | 7 % (n=293) | 6 % (n=169) | 5 % (n=2,384) |
| 1-2 days | 23 % (n=900) | 24 % (n=706) | 31 % (n=13,547) |
| 3-4 days | 22 % (n=878) | 25 % (n=711) | 26 % (n=11,412) |
| 5-6 days | 15 % (n=608) | 13 % (n=370) | 13 % (n=5,647) |
| 7 days | 31 % (n=1,226) | 31 % (n=901) | 22 % (n=9,615) |
| Missing | 1 % (n=56) | 2 % (n=45) | 2 % (n=944) |
| **Fizzy Drinks (How many days did you drink at least one fizzy drink [e.g. coke, fanta, sprite]?)** |  |  |  |
| 0 days | 27 % (n=1,071) | 30 % (n=869) | 37 % (n=16,165) |
| 1-2 days | 35 % (n=1,367) | 34 % (n=999) | 38 % (n=16,413) |
| 3-4 days | 14 % (n=573) | 15 % (n=427) | 12 % (n=5,428) |
| 5-6 days | 6 % (n=245) | 7 % (n=190) | 4 % (n=1,806) |
| 7 days | 16 % (n=649) | 12 % (n=359) | 6 % (n=2,800) |
| Missing | 1 % (n=56) | 2 % (n=58) | 2 % (n=937) |
| **Takeaways (How many days did you eat take away foods [e.g. McDonalds, KFC, Chinese]?)** |  |  |  |
| 0 days | 35 % (n=1,387) | 39 % (n=1,126) | 44 % (n=19,349) |
| 1-2 days | 43 % (n=1,722) | 42 % (n=1,229) | 44 % (n=19,150) |
| 3-4 days | 10 % (n=392) | 9 % (n=248) | 6 % (n=2,590) |
| 5-6 days | 4 % (n=142) | 4 % (n=108) | 2 % (n=760) |
| 7 days | 6 % (n=233) | 5 % (n=150) | 2 % (n=893) |
| Missing | 2 % (n=85) | 1 % (n=41) | 2 % (n=807) |
| **Physical Activity (How many days were you physically active [got out of breath]?)** |  |  |  |
| 0 days | 10 % (n=405) | 7 % (n=213) | 6 % (n=2,512) |
| 1-2 days | 26 % (n=1,038) | 27 % (n=797) | 28 % (n=11,988) |
| 3-4 days | 23 % (n=910) | 26 % (n=748) | 27 % (n=11,729) |
| 5-6 days | 17 % (n=654) | 16 % (n=464) | 19 % (n=8,280) |
| 7 days | 23 % (n=917) | 22 % (n=632) | 19 % (n=8,367) |
| Missing | 1 % (n=37) | 2 % (n=48) | 2 % (n=673) |
| **Sleep (Sleep hours calculated from: What did you wake up today and What time did you go to sleep?)** |  |  |  |
| 0-3 hours | 3 % (n=105) | 1 % (n=40) | 1 % (n=201) |
| 4-6 hours | 11 % (n=440) | 7 % (n=196) | 3 % (n=1,304) |
| 7-9 hours | 44 % (n=1,742) | 41 % (n=1,202) | 33 % (n=14,556) |
| 9-12 hours | 35 % (n=1,370) | 42 % (n=1,228) | 54 % (n=23,320) |
| 12+ hours | 6 % (n=238) | 6 % (n=183) | 7 % (n=3,225) |
| Missing | 2 % (n=66) | 2 % (n=53) | 1 % (n=943) |

Based on the demographic data for the study population, the sample is distributed nearly evenly between boys and girls, with a smaller proportion identifying as Prefer Not To Say. Boys and the Prefer Not To Say group report higher levels of behavioural difficulties across the cohort. Clinical behavioural difficulties are higher in screen time use between 5-6 days, 7 days of sugary snacks, fizzy drinks and takeaways, 0 days of physical activity and 0-3 hours sleep.

### Regression Analysis

A multivariable linear regression model (table 2) was fitted with behavioural difficulty score as the outcome variable. Predictor variables included gender, ALN status, 7 days a week screen time, sugary snack consumption (<4 days associated with lower difficulties), fizzy drink consumption, takeaway consumption, physical activity, and sleep duration. This approach enabled an exploration of the association between each health behaviour and behavioural difficulties while adjusting for the potential influence of the other behavioural and demographic factors. A coefficient of 0.48 for boys indicates that boys had, on average, a 0.48-point higher behavioural difficulty score than girls, after adjustment for the other variables in the model.

**Table 2.** Linear regression model examining the association between behavioural difficulty score and health behaviours (n=50412)

|  | Coeff. | Std. Err. | p>t | Upper | Lower |
| --- | --- | --- | --- | --- | --- |
| **Gender** |  |  |  |  |  |
| Girl | Ref. | Ref. | Ref. | Ref. | Ref. |
| Boy | 0.48 | 0.02 | 0.00 | 0.44 | 0.52 |
| Prefer | 0.79 | 0.07 | 0.00 | 0.66 | 0.93 |
| **ALN Status** |  |  |  |  |  |
| No ALN | Ref. | Ref. | Ref. | Ref. | Ref. |
| ALN | 0.33 | 0.02 | 0.00 | 0.29 | 0.38 |
| **Screen Time** |  |  |  |  |  |
| 0 days | Ref. | Ref. | Ref. | Ref. | Ref. |
| 1-2 days | -0.02 | 0.06 | 0.70 | -0.13 | 0.09 |
| 3-4 days | -0.08 | 0.06 | 0.15 | -0.19 | 0.03 |
| 5-6 days | 0.01 | 0.06 | 0.85 | -0.10 | 0.12 |
| 7 days | 0.27 | 0.05 | 0.00 | 0.16 | 0.37 |
| **Sugary Snacks** |  |  |  |  |  |
| 0 days | Ref. | Ref. | Ref. | Ref. | Ref. |
| 1-2 days | -0.23 | 0.05 | 0.00 | -0.32 | -0.14 |
| 3-4 days | -0.17 | 0.05 | 0.00 | -0.26 | -0.07 |
| 5-6 days | -0.03 | 0.05 | 0.56 | -0.13 | 0.07 |
| 7 days | 0.05 | 0.05 | 0.34 | -0.05 | 0.15 |
| **Fizzy Drinks** |  |  |  |  |  |
| 0 days | Ref. | Ref. | Ref. | Ref. | Ref. |
| 1-2 days | 0.15 | 0.02 | 0.00 | 0.10 | 0.20 |
| 3-4 days | 0.22 | 0.03 | 0.00 | 0.15 | 0.28 |
| 5-6 days | 0.30 | 0.05 | 0.00 | 0.19 | 0.40 |
| 7 days | 0.57 | 0.04 | 0.00 | 0.49 | 0.66 |
| **Takeaways** |  |  |  |  |  |
| 0 days | Ref. | Ref. | Ref. | Ref. | Ref. |
| 1-2 days | 0.07 | 0.02 | 0.00 | 0.03 | 0.11 |
| 3-4 days | 0.40 | 0.04 | 0.00 | 0.31 | 0.49 |
| 5-6 days | 0.44 | 0.07 | 0.00 | 0.30 | 0.59 |
| 7 days | 0.56 | 0.07 | 0.00 | 0.42 | 0.70 |
| **Physical Activity** |  |  |  |  |  |
| 0 days | Ref. | Ref. | Ref. | Ref. | Ref. |
| 1-2 days | -0.25 | 0.05 | 0.00 | -0.34 | -0.16 |
| 3-4 days | -0.30 | 0.05 | 0.00 | -0.40 | -0.21 |
| 5-6 days | -0.39 | 0.05 | 0.00 | -0.49 | -0.30 |
| 7 days | -0.23 | 0.05 | 0.00 | -0.33 | -0.14 |
| **Sleep** |  |  |  |  |  |
| Sleep hours | -0.19 | 0.01 | 0.00 | -0.20 | -0.17 |

Boys showed significantly higher behavioural difficulty scores than girls (Coeff. 0.48, 95 % CI: 0.44-0.52, p<0.001) and children with ALN also reported higher scores (0.33, 95 % CI: 0.29-0.38, p<0.001). Among health behaviours, daily consumption of fizzy drinks (0.57, 95 % CI: 0.49-0.66. p<0.001) and takeaway food (0.56, 95 % CI: 0.42-0.70, p<0.001) were associated with the largest increases in behavioural difficulty scores. A higher behavioural difficulty score was associated with increasing frequency of consumption.

Higher levels of physical activity and longer sleep duration (-0.19, 95 % CI: -0.20—0.17, p<0.001) were associated with lower behavioural difficulty scores. Reporting at least 60 minutes of physical activity on multiple days per week consistently demonstrated lower behavioural difficulty scores, particularly in the 5-6 days group (-0.39, 95 % CI: -0.49—0.30, p<0.001). Screen time was associated with behavioural difficulties only at the highest level, with children reporting two or more hours of screen time on all seven days of the week showing significantly higher behavioural difficulty scores (0.27, 95 % CI: 0.16-0.37, p<0.001).

These findings suggest that lower sleep duration, frequent consumption of fizzy drinks and takeaway foods, and consistently high screen time are independently associated with increased behavioural difficulties in primary school-aged children. There is also evidence that higher physical activity can be protective. These findings remained after accounting for gender and ALN status, indicating that health behaviours may provide explanatory value beyond established demographic and behavioural factors.

### Cluster Analysis Of Health Behaviours

To explore the associations between screen time, diet, and behavioural difficulty, a K-means cluster analysis was conducted, identifying four distinct behavioural profiles within the cohort. This approach was selected to identify how health behaviours can co-exist to provide a view of how child health can cluster. The four clusters represent 32 %, 18 %, 24 %, and 26 % of the population, respectively, with each group displaying a unique profile. Physical activity and sleep were also included in the analysis as potential mediators.

**Table 2.** Cluster analysis of health behaviours.

| Cluster | Behaviour | Screen Time | Sugary Snacks | Fizzy Drinks | Takeaways | Physical Activity | Sleep Hours | % Of Cohort | % Of ALN Cohort |
| --- | --- | --- | --- | --- | --- | --- | --- | --- | --- |
| Low behavioural difficulties, high screen time, good sleep | -0.34 | 0.73 | 0.46 | -0.33 | -0.21 | -0.09 | 0.22 | 32 | 28 |
| *Std. Dev.* | *0.69* | *0.48* | *0.91* | *0.60* | *0.68* | *0.98* | *0.71* |  |  |
| High behaviour difficulties, high fizzy drinks, poor sleep | 0.30 | 0.57 | 0.75 | 1.69 | 1.03 | -0.10 | -0.39 | 15 | 20 |
| *Std. Dev.* | *1.02* | *0.77* | *0.88* | *0.88* | *1.40* | *1.07* | *1.17* |  |  |
| Low behavioural difficulties, low screen time, good sleep | -0.37 | -0.96 | -0.55 | -0.39 | -0.27 | 0.07 | 0.33 | 37 | 33 |
| *Std. Dev.* | *0.70* | *0.59* | *0.74* | *0.61* | *0.75* | *0.95* | *0.73* |  |  |
| Very high behavioural difficulties, poor sleep | 1.25 | 0.28 | -0.27 | -0.13 | -0.02 | 0.08 | -0.85 | 16 | 19 |
| *Std. Dev.* | *0.96* | *0.87* | *0.92* | *0.72* | *0.89* | *1.02* | *1.22* |  |  |

In Cluster 1: Low behavioural difficulties, high screen time (32 %), fizzy drink and takeaway consumption are below average, and they have good sleep hygiene but report the highest levels of screen time (0.73). Despite this, Cluster 1 has lower behavioural scores (-0.34), indicating diet and sleep is associated with behavioural difficulties.

Cluster 3: Low behavioural difficulties, low screen time (37 % of the population), represents the group with the lowest behavioural difficulties. This group is characterised by lower-than-average levels of screen time (-0.98), sugary snack intake (-0.55), and fizzy drink consumption (-0.39) and slightly higher physical activity and sleep duration.

In contrast, Cluster 4: Very high behavioural difficulties, poor sleep (16 %) has the highest behavioural difficulty score with the least sleep, alongside high screen time.

The remaining cluster suggests there are strong dietary associations with behavioural difficulties. Cluster 2: High behavioural difficulties, high fizzy drinks: (15 %) consumes high levels of fizzy drinks (1.69) and takeaways (1.03), coupled with the poor sleep (-0.39) and the behavioural difficulties (0.30).

### Longitudinal Trends in Health Behaviours

Trends in the data (Figure 1) show the percentage of children reporting screen time has increased over the years, with nearly half of the cohort (42 %) reporting engaging with a screen for more than two hours across 7 days of the week. Similar trends are seen in sugary snacks and fizzy drinks, with annual increases and a peak in the year 2021. Children appear to be snacking more regularly than a decade ago.

**Figure 1.**
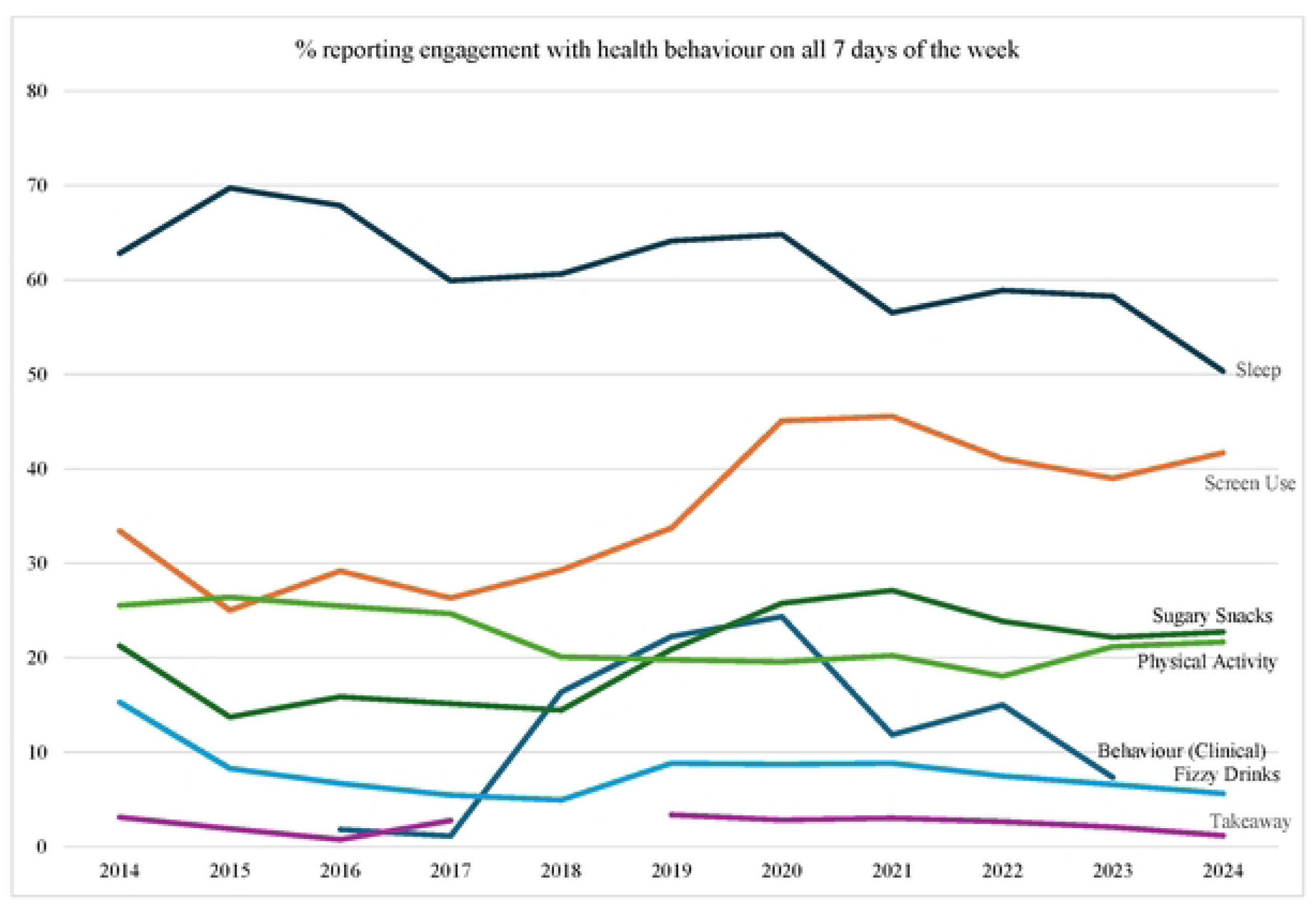
Annual Trends in Children’s Self-Reported Health Behaviours (n=55966)

Over the last decade, children’s health behaviours show some improvements, but these improvements may be offset by rising screen time and plateauing physical activity. The post-pandemic period appears to be associated with a lasting increase in daily screen use that has not returned to pre-pandemic levels. Figure 1 suggests that daily screen use increased substantially over the study period, particularly between 2019 and 2021, and remained elevated.

Behavioural difficulties increased sharply during the pandemic period before falling, whereas sleep duration showed a decline. Together, these findings suggest that screen-related behaviours experienced the greatest increase over the decade, while physical activity remained comparatively stable. A notable increase occurred between 2019 and 2020, likely reflecting the disruption caused by the COVID-19 pandemic. Screen time saw its sharpest increase during this window, peaking in 2020, and remained elevated well above pre-2019 levels through to 2024. While most behaviours surged during the pandemic, takeaway consumption remained relatively stable. Despite a public health emphasis on physical activity, there is little evidence of a sustained increase in activity levels in children.

## Discussion

This study examined the relationship between demographic characteristics, daily health behaviours, and behavioural difficulties. Multivariable regression analysis demonstrated that being a boy, having an ALN diagnosis and the daily consumption of fizzy drinks and takeaway foods showed the strongest independent associations with higher behavioural difficulties. The results relating to ALN suggest behaviour may also be an important equity and intervention issue. It would be useful to reflect on whether children with ALN may experience different barriers to sleep, diet, physical activity and screen use, and whether universal school-based approaches need to be complemented by more tailored support. This would help avoid presenting ALN only as a risk factor and instead frame it as a marker of where more inclusive, needs-led support may be required.

These findings were further clarified by cluster analysis, which identified four distinct behavioural profiles. Notably, children in clusters characterized by low behavioural difficulties maintained good sleep hygiene and below-average dietary intake of fizzy drinks and takeaways, even when displaying higher levels of daily screen time (Cluster 1). The highest behavioural difficulty scores clustered with poor sleep, high screen use, and poor dietary habits (Clusters 2 and 4). Finally, longitudinal trends over the decade revealed a post-pandemic rise in daily screen time and a gradual decline in sleep duration, whilst physical activity levels remained plateaued.

The results indicate that although high sugar snacking and screen time have often been linked with behaviour (6,10,14,23), other health behaviours appear to have a greater association. Specifically, these findings show that sleep duration and diet (particularly fizzy drinks and takeaways) are associated with behavioural difficulties in children. This suggests that how much sleep children are having coupled with what they are eating and drinking has significant implications for behaviour. Insufficient sleep and sleep quality have been shown to affect cognitive function in children, including attention and academic performance (27). Previous research has shown links between sleep duration and acting out behaviours and behavioural difficulties (28,29). Therefore, given the evidence presented here, it is important that settings promote healthy sleep habits. This could include school education, parental intervention or broader public health campaigns. Wider evidence is showing an increasing prevalence in child sleep problems (27). The impact of screen time finding is interesting because high screen time does not appear uniformly harmful across clusters, especially where sleep and diet profiles are more favourable. It may suggest that total screen time alone is too blunt a measure, and that the context, timing, content and displacement of sleep or activity may be more important than duration alone.

Sleep also moderates the relationship between screen use and behaviour (14). These findings contribute to a growing body of evidence suggesting that how children spend their time, especially how much sleep they have, diet such as take aways and fizzy drinks, may have implications for behaviour. Diet quality has been shown to influence mood, attention, and executive functioning, potentially through fluctuations in blood sugar (23). This has implications for school food culture, (37) particularly when considering food-based policy and implementation such as the universal provision of free school meals (UPFSM) (38) in primary education in Wales. This means that focus should be paid to the quality of food being offered to children; prioritising less processed food. Recent qualitative findings from children and parents reveal significant concerns around the food provided to children. Children report meal quality had a direct impact on their mood and their ability to concentrate, whilst parents highlighted that school meals contain a significant amount of ultra-processed foods. These observations are concerning given the findings in the present study noting that ultra-processed food consumption is associated with behavioural difficulties. This concern is supported by recent evidence demonstrating that ultra-processed food consumption in early childhood is associated with adverse behavioural and emotional outcomes (38) as well as negative impacts on children’s mental health (39), emphasising the need for substantial improvements in school meal quality. However, only so much of this can be done via school-based intervention.

There is now guidance being implemented around 24-hour behaviours (including sleep) rather than focussing on each behaviour in isolation (40). These challenges overly simplistic messages that focus solely on doing more or less of one behaviour and instead support approaches that address the broader behaviours that children can engage with across the day. When schooling increasingly utilising technology and screens for educational purposes during the day, safeguarding the remaining hours of a child’s day for adequate sleep and proper nutrition becomes even more vital. The results support a shift away from single-behaviour messaging, such as reducing screen time alone, towards a more integrated 24-hour behaviour approach that includes sleep, diet, movement and screen use together. This could be of real value for schools and policymakers.

Current recommendations often emphasise limiting total screen time (e.g. two hours per day). This work suggests that greater emphasis should be placed on sleep with additional focus on food choices and embedding opportunities for regular movement throughout the day. For schools, this supports approaches to consider behaviours across the 24-hour period and focussing less on improving individual behaviours and more on promoting a healthy balance across a range of behaviours. This would include education that can be taken home to influence dietary choices and sleep outside of school. Given the observed associations with behavioural difficulties, such strategies may offer wider benefits for classroom behaviour, attention, and learning. In Wales, these approaches align well with the Curriculum for Wales, where Health and Wellbeing is a core Area of Learning Experience (31).

This also reinforces the idea that public health interventions might be less about increasing physical activity in isolation and more about helping to promote a balanced lifestyle. This is important when coupled with the finding that trends in physical activity have remained stagnant or have declined. This finding is consistent with previous work highlighting the importance of different behaviours (physical activity, sedentary behaviour, and sleep) across the day (8,41). The longitudinal trends observed from 2014 to 2024 demonstrate a stable and, in some areas, improving levels of health behaviours across Wales. This is important to note as behavioural difficulties and sleep both seem to be improving.

Analysis by gender and ALN status revealed some disparities, with boys consistently reporting higher mean scores for behavioural difficulties than girls as well as those with ALN highlighting an area of inequality for support.

### Limitations

This study has several limitations that should be considered when interpreting the findings. Measures were self-reported, which may introduce reporting bias, including recall inaccuracies. Children may under- or over-estimate their screen time, dietary behaviours, and physical activity, which could influence the observed associations. Screen time was measured as total time and did not distinguish between different types of screen use. Therefore, was not possible to differentiate between potentially beneficial activities, such as educational use, and more passive forms of screen use. This limits the ability to draw conclusions about how specific types of screen-based behaviours may differentially relate to behavioural outcomes. Finally, while this study focused on key health behaviours, it did not account for a range of additional influences that may shape children’s behaviour.

## Conclusions

This study highlights the association between screen time, sugary snack consumption, fizzy drinks, takeaways, physical activity, sleep duration and behavioural difficulties in school-aged children. Children who have poor dietary behaviours and poor sleep time showed the greatest behavioural difficulties regardless of ALN status, reinforcing the importance of integrating public health education and provision across a variety of health areas. It is worth noting that these behaviours are shaped by wider home, school and community environments, rather than being solely individual or parental choices. There is a public health framing around supportive environments, school food quality, access to physical activity, sleep routines and wider inequalities.

The key priority is to move beyond single behaviour guidance that targets solitary health aspects in isolation. Public health messaging must evolve to address the clustering of high-risk dietary intake (specifically ultra-processed options like takeaways and fizzy drinks) and sleep habits. As well as acknowledging that physical activity can mitigate screen use. For example, improving children’s sleep behaviours such as clear bedtime, reducing take aways and fizzy drinks and processed food may be a practical and impactful ways to support improved behaviour.

Ultimately, supporting families and schools to have more balanced 24-hour behaviour profiles will provide the most effective foundation for improving behavioural difficulties. Future research should consider a broader set of contextual and behavioural factors, including different types of movement behaviours, more detailed dietary patterns such as types of snacking, the nature and context of technology use and the content of screen engagement. Incorporating these factors would provide a more comprehensive understanding of the pathways linking these behaviours and behavioural difficulties.

## Data Availability

The HAPPEN Cohort dataset is available in the Secure Anonymised Information Linkage (SAIL) Databank upon reasonable request.

## Acknowledgements

The HAPPEN team would like to acknowledge the pupils, teachers and schools who are a part of HAPPEN. Without them, this research would not be possible.

